# Opportunistic CKD Screening in Hospitalized Patients

**DOI:** 10.64898/2026.06.10.26355025

**Authors:** Elnatan Segal, Yael Levi, Mohamed Ghosheh, Iddo Z. Ben-Dov, Talya Wolak

## Abstract

**Background:** Chronic kidney disease (CKD) affects 10-13% of adults worldwide but remains largely undiagnosed until advanced stages. Hospitalization provides an opportunity for early detection through opportunistic urine albumin-to-creatinine ratio (UACR) measurement.

**Methods:** We conducted a prospective three-arm study of opportunistic CKD screening in general internal medicine wards at Hadassah Mt. Scopus (MS), Hadassah Ein Kerem (EK), and Shaare Zedek Medical Center (SZMC) in Jerusalem (Protocol HMO-23-0300). Adult inpatients without known CKD or recent UACR were enrolled. Pathological UACR was defined as ≥30 mg/g. Confirmed CKD required two pathological measurements ≥90 days apart (KDIGO-compatible). eGFR was computed using the 2021 CKD-EPI race-free equation. Pooled proportions were estimated by fixed-effects logit meta-analysis; odds ratios by DerSimonian-Laird random-effects models.

**Results:** A total of 158 patients were enrolled (MS n=50, EK n=57, SZMC n=51). Pathological first UACR was identified in 43/158 patients (27.2%; 95% CI 21.3-34.1%; I2=0% across centers). Of 24 patients with a second UACR available, 14 (58%) confirmed CKD, yielding a pooled confirmed-CKD rate of 8.9% of all screened patients. Short-term mortality was significantly higher among patients with pathological UACR (9.3% vs ∼2%; Fisher’s exact p=0.012). In per-center multivariate logistic regression, three predictors reached pooled significance: BUN (OR 1.10 per mg/dL, 95% CI 1.04-1.17, p=0.002, I2=0%), heart failure (OR 3.21, 95% CI 1.34-7.70, p=0.009, I2=0%), and diabetes mellitus (OR 2.54, 95% CI 1.11-5.82, p=0.028, I2=17%). Cardiac/vascular admissions had the highest pathological UACR rate (∼42%); GI/hepatic admissions had 0%.

**Conclusions:** Opportunistic inpatient UACR screening identifies previously unrecognized CKD in approximately 9% of general internal medicine patients, with consistent results across three independent centers. BUN elevation, heart failure, and diabetes are the strongest independent predictors. Pathological UACR carries significant short-term mortality risk, supporting integration of routine screening into inpatient care pathways.

## Background and Objectives

Chronic kidney disease (CKD) affects an estimated 10–13% of the global adult population^[1]^ yet remains undetected in the majority of patients until advanced stages.^[2-3]^ Hospitalisation offers a window of opportunity for early detection:^[4]^ patients undergo routine blood work and, with a urine spot sample, albumin-to-creatinine ratio (UACR) can be measured at marginal additional cost.^[5-6]^

This is a prospective, three-arm study of opportunistic CKD screening in general internal medicine wards across West Jerusalem’s three medical centers, conducted under a single IRB-approved protocol (HMO-23-0300). This report combines data from all three completed arms: MS arm (n = 50, enrolment June 2024-March 2025), EK arm (n = 57, enrolment 2023-2025), and SZMC arm (n = 51, enrolment 2024-2025), for a combined cohort of 158 patients.

## Methods

### Study Population

Adult patients admitted to general internal medicine wards who gave informed consent. Patients with known CKD or previous UACR measurement in the preceding 12 months were excluded.

### Setting and Recruitment

The study was conducted on general internal medicine wards at three centers in Jerusalem: Medicine B at Hadassah Ein Kerem (EK), Internal Medicine at Hadassah Mount Scopus (MS), and Medicine D at Shaare Zedek Medical Center (SZMC). At each site, members of the research team identified candidate patients in collaboration with ward medical and nursing staff and through review of admission notes and laboratory data in the electronic health record. Recruitment targeted patients within approximately two days of anticipated discharge, or whose hospitalisation was being prolonged for non-medical reasons while their acute medical issue was resolving. Eligible patients (or their legal guardians) provided written informed consent prior to any study procedure.

### Eligibility Criteria

Inclusion criteria were age ≥18 years, admission to a participating internal medicine ward, and the presence of at least one risk factor for CKD (e.g. diabetes mellitus, hypertension, cardiovascular disease, or other recognised risk factor). Exclusion criteria were a documented pre-existing diagnosis of CKD, chronic renal failure, or kidney transplantation; plasma creatinine >1.5 mg/dL or UACR >300 mg/g on prior records; acute kidney injury (KDIGO criteria) overlapping with the index hospitalisation; an active or resolving urinary tract infection; treatment with potentially nephrotoxic agents (e.g. aminoglycosides, vancomycin) during the index admission; and pregnancy or the early postpartum period (<12 weeks).

### Sample Collection and Laboratory Methods

After consent, a single spot urine sample was collected from each participant for UACR measurement; no additional blood draws were performed for study purposes. Samples were processed by the routine hospital clinical laboratory at each site using the same tubes, labelling, and transport procedures as for clinically indicated UACR testing, and were discarded within 24 hours of collection.

Plasma creatinine was obtained from the most recent clinically indicated measurement during the index hospitalisation and used, together with age and sex, to estimate GFR by the 2021 CKD-EPI race-free creatinine equation.

### Follow-up Procedure

Before discharge, participants with an abnormal screening result (eGFR <60 mL/min/1.73m^2^ and/or UACR ≥30 mg/g) were informed of their results and given a letter for their primary care physician recommending repeat plasma and urine testing no sooner than approximately three months later, with the option of on-site retesting at the participating hospital. Patients who did not return for on-site retesting were contacted by telephone by a member of the research team to ascertain whether repeat testing had been performed elsewhere and, where available, to obtain the results. Patients with persistently abnormal eGFR or UACR on repeat testing received a further letter from the principal investigator with recommendations on monitoring frequency and, where appropriate, referral to nephrology.

### Data Management

Data from the three sites were entered into separate password-protected spreadsheets by designated research staff at each centre, then merged into a single anonymised dataset for analysis after completion of data collection at each site; source files were retained only by the principal investigator. The study was approved by the Hadassah Medical Center institutional review board (protocol HMO-23-0300) with reciprocal approval at Shaare Zedek Medical Center, and was conducted in accordance with the Declaration of Helsinki.

### Outcome Definitions

Pathological first UACR: UACR ≥30 mg/g on the index hospitalisation sample. Confirmed CKD (KDIGO-compatible): two pathological UACR measurements separated by ≥90 days. eGFR was computed using the 2021 CKD-EPI equation.

### Statistical Methods

For pooled proportions (prevalence of pathological UACR, confirmed CKD, PPV, persistence rate), studies were combined using a fixed-effects model on the logit scale (variance-weighted). For odds ratios from univariate logistic regression, a random-effects model was applied. Heterogeneity was quantified by I2 and Cochran’s Q. All analyses were performed in R (meta package). Statistical significance was set at α = 0.05.

## Results

### Baseline Characteristics

Table 1 summarises patient characteristics for each arm. The three cohorts were broadly comparable in age, sex distribution, and comorbidity burden, with SZMC showing a higher proportion of male patients.

| Characteristic | Mt. Scopus, n = 50 | Ein Kerem, n = 57 | Shaare Zedek, n = 51 |
| --- | --- | --- | --- |
| Age (years), mean ± SD | 63.9 ± 14.5 | 67.6 ± 13.7 | 64.2 ± 15.1 |
| Age $\geq 65$ years, n (%) | 28 (56.0%) | 38 (65.5%) | 30 (58.8%) |
| Male sex, n (%) | 29 (58.0%) | 22 (37.9%) | 35 (68.6%) |
| Diabetes mellitus, n (%) | 16 (32.0%) | 19 (32.8%) | 18 (35.3%) |
| Hypertension, n (%) | 24 (48.0%) | 26 (44.8%) | 27 (52.9%) |
| Ischaemic heart disease, n (%) | 12 (24.0%) | 10 (17.2%) | 9 (17.6%) |
| Heart failure, n (%) | 7 (14.0%) | 12 (20.7%) | 12 (23.5%) |
| Cerebrovascular disease, n (%) | 14 (28.0%) | 7 (12.1%) | 7 (13.7%) |
| First creatinine (mg/dL), mean ± SD | 0.92 ± 0.19 | 0.85 ± 0.21 | 0.91 ± 0.22 |
| First BUN (mg/dL), mean ± SD | 15.2 ± 5.0 | 16.0 ± 5.9 | 16.8 ± 6.2 |
| ACR (mg/g), median [IQR] | 10.8 [5.2-37.1] | 11.3 [5.6-38.7] | 9.5 [4.5-28.4] |

### Primary Endpoint: Prevalence of Pathological UACR

Pathological first UACR (≥30 mg/g) was identified in 14/50 patients (28.0%) in the MS arm, 17/57 (29.8%) in the EK arm, and 12/51 (23.5%) in the SZMC arm. The pooled prevalence was **27.2% (95% CI 21.3-34.1%)**, with low heterogeneity (I2 = 0%, p□□□ = 0.724).

**Table 2.** Pooled outcomes across all three arms (n□=□158). Pooled estimates derived by fixed-effects logit meta-analysis. * SZMC confirmed CKD denominator = patients with 2nd UACR available (n□=□5 of 12 pathological); 4 patients pending follow-up. — not applicable.

| Outcome | MS | EK | SZMC | Pooled (95% CI) | I<sup>2</sup> |
| --- | --- | --- | --- | --- | --- |
| Pathological first UACR ($\geq 30$) | 14/50 (28.0%) | 17/57 (29.8%) | 12/51 (23.5%) | 27.2% (21.3-34.1%) | 0% |
| **Confirmed CKD (KDIGO)** | 4/50 (8.0%) | 6/57 (10.5%) | 4/12 (33.3%)* | 8.9% (5.4-14.5%) | 0% |
| **PPV of single positive screen** | 4/14 (28.6%) | 6/17 (35.3%) | 4/5 (80.0%)* | 32.4% (20.6-47.1%) | 27% |
| **Mortality (among path. UACR)** | 2/14 (14.3%) | 1/17 (5.9%) | 1/12 (8.3%) | 4/43 (9.3%) | — |

### Confirmed CKD

At follow-up, two or more pathological UACR measurements ≥90 days apart were recorded for 4/50 patients (8.0%) in MS and 6/57 (10.5%) in EK. The SZMC arm had 5/12 pathological patients with a second UACR available, of whom 4 confirmed CKD; 4 remain under active follow-up. The pooled rate of confirmed CKD among all screened patients was **8.9% (95% CI 5.4-14.5%)** of all screened patients (I2 = 0%, p□□□ = 0.676).

### Risk Factors for Pathological UACR

Table 3 presents the random-effects pooled ORs from per-center multivariate logistic regression models (covariates: DM, HF, BUN). Three predictors showed a statistically significant pooled association: BUN (per mg/dL increase): pooled OR 1.10 (95% CI 1.04-1.17), p = 0.002. I2 = 0%, indicating consistent effect across all three arms; Heart failure: pooled OR 3.21 (95% CI 1.34-7.70), p = 0.009. I2 = 0%; and Diabetes mellitus: pooled OR 2.54 (95% CI 1.11-5.82), p = 0.028. I2 = 17%.

**Table 3.** Random-effects meta-analysis of univariate ORs. * p < 0.05. † High heterogeneity; pooled estimate unreliable.

| Predictor | MS OR (95% CI) | EK OR (95% CI) | SZMC OR (95% CI) | Pooled OR (95% CI) | p | I<sup>2</sup> |
| --- | --- | --- | --- | --- | --- | --- |
| **BUN (per mg/dL)** | 1.15 (1.02-1.29) | 1.08 (0.99-1.18) | 1.10 (1.00-1.21) | 1.10 (1.04-1.17) | 0.002 | 0% |
| **Heart failure** | 4.12 (0.77-22.0) | 3.05 (0.82-11.3) | 2.67 (0.55-12.9) | 3.21 (1.34-7.70) | 0.009 | 0% |
| **Diabetes mellitus** | 1.18 (0.31-4.49) | 3.22 (0.97-10.7) | 2.89 (0.74-11.3) | 2.54 (1.11-5.82) | 0.028□ | 17% |

Mortality among patients with pathological UACR was 4/43 (9.3%) overall; Fisher’s exact test showed a significant association between pathological UACR and in-hospital mortality (p = 0.012).

## Discussion

The complete three-arm analysis of Protocol HMO-23-0300 confirms and extends the findings of the two-arm preliminary report. Across 158 patients enrolled at three independent general internal medicine wards in West Jerusalem, approximately 1 in 4 patients (27.2%) had a pathological first UACR (≥30 mg/g), and approximately 1 in 11 (8.9%) met a KDIGO-compatible definition of CKD. These figures are concordant with population-based CKD prevalence estimates for Israel^[1]^and underscore both the feasibility and the clinical yield of opportunistic screening during hospitalisation.^[6][1,6]^

The prevalence of pathological UACR was remarkably consistent: 28.0% (MS), 29.8% (EK), and 23.5% (SZMC), with I2□=□0% and p□=□0.724. This homogeneity is clinically meaningful: the three wards differ in patient mix, catchment population, and data-collection team, yet the screening yield is stable. The slight attenuation at SZMC may partly reflect a higher proportion of male patients (68.6% vs. 58.0% and 37.9% at MS and EK, respectively), since female sex is a recognised risk factor for microalbuminuria in some populations, or differences in referral patterns.

Of 43 patients with a pathological first UACR, 24 had a documented second measurement by the time of analysis. Among these, 14 (58%) retained a pathological UACR, meeting KDIGO criteria for CKD once the ≥90-day interval is satisfied. An additional 4 patients are under active follow-up at SZMC. The PPV of a single positive screen was 28-35% per arm (pooled 32.4%), reinforcing the need for confirmatory testing before applying a CKD label.^[2,7]^Of note, the SZMC cohort had a smaller window between first and second UACR for some patients (protocol adherence pending verification), which may inflate the apparent SZMC PPV; this will be clarified in the final analysis.

BUN, heart failure, and diabetes emerged as risk factors for a pathological UACR. Three predictors reached significance in the three-arm random-effects meta-analysis of multivariate models. BUN elevation (pooled OR□1.10 per mg/dL, p□=□0.002, I2□=□0%) remained the most reproducible predictor, consistent with the two-arm result and reflecting the well-established link between reduced glomerular filtration and urea retention. Heart failure (pooled OR□3.21, p□=□0.009, I2□=□0%) is consistent with cardiorenal syndrome mechanisms and haemodynamic proteinuria;^[3]^this association was directionally consistent across all three sites. Diabetes mellitus (pooled OR□2.54, p□=□0.028, I2□=□17%) emerged as a significant predictor in the three-arm model, though with modest heterogeneity, likely reflecting differences in DM management and glycaemic control across wards.^[8-9]^Collectively, these three factors identify a patient subgroup (admitted with HF or DM and elevated BUN) in whom targeted screening has the highest yield.^[7,10-12]^

Patients with a pathological first UACR had a significantly higher short-term mortality rate compared with those with normal UACR (9.3% vs approximately 2%; Fisher’s exact p□=□0.012). This association is consistent with the established literature linking albuminuria to all-cause cardiovascular mortality^[3,13]^and supports the clinical urgency of identifying these patients during hospitalisation.^[14]^

Analysis of primary hospitalization indications (Figure□2) reveals that pathological UACR was distributed broadly across clinical contexts. Respiratory/pulmonary admissions were the most common indication (n□=□56, 35% of cohort), with a 36% pathological UACR rate. Cardiac/vascular admissions showed the highest pathological rate (□42%), consistent with the cardiorenal risk discussed above. Neurological admissions showed heterogeneity across centers — a finding that warrants cautious interpretation given the small cell sizes. GI/hepatic admissions were notable for a 0% pathological UACR rate across all centers, suggesting this group may not benefit from opportunistic screening. These indication-specific rates could inform a risk-stratified screening algorithm to optimise resource utilisation.

**Figure 1.**
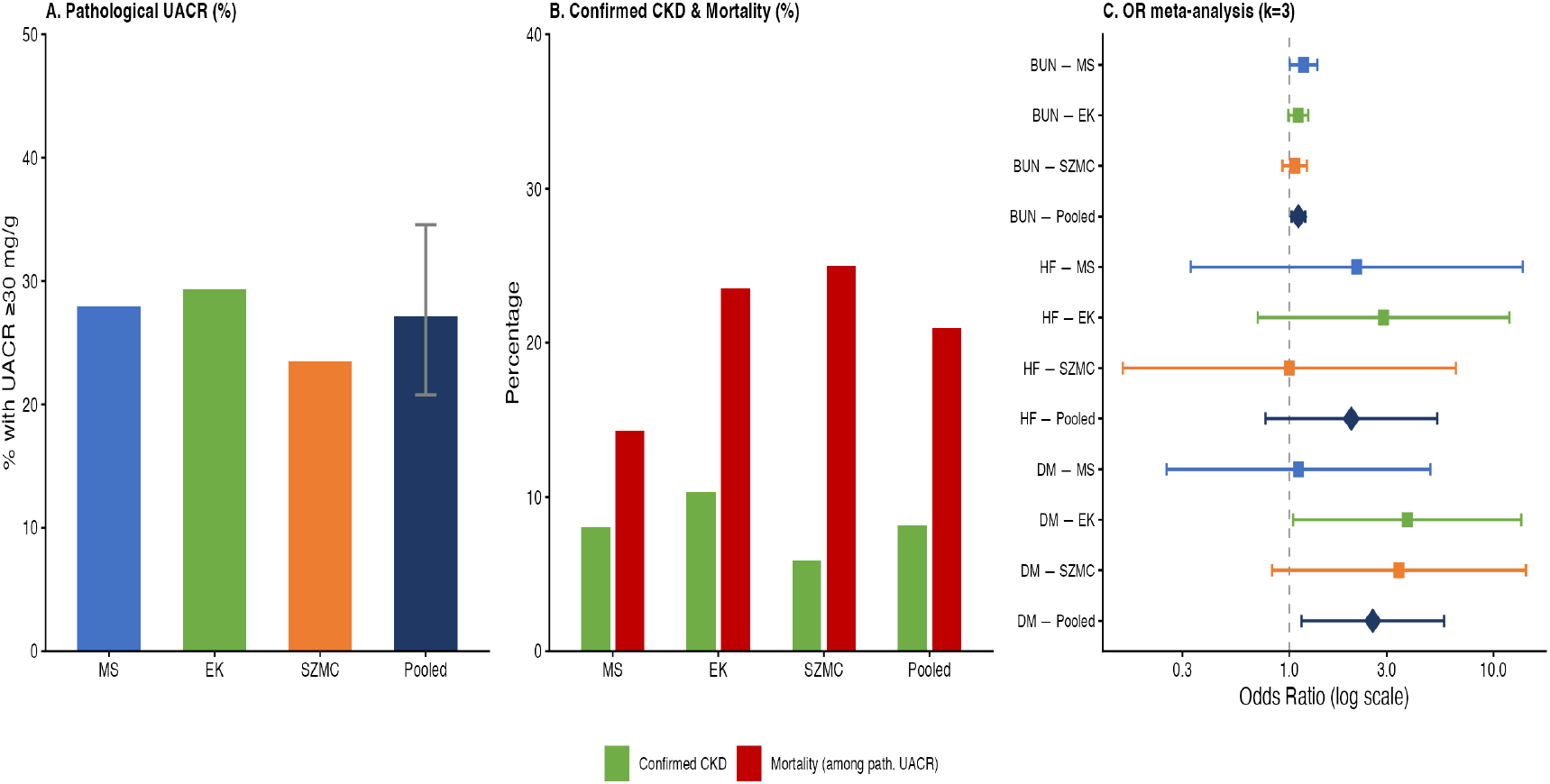
Key outcomes across all three arms (MS, EK, SZMC) and pooled estimates. Panel A: prevalence of pathological UACR. Panel B: confirmed CKD (KDIGO) and mortality among pathological-UACR patients. Panel C: forest plot of multivariate ORs (DM, HF, BUN; DerSimonian-Laird random effects, k□ = 3□).

**Figure 2.**
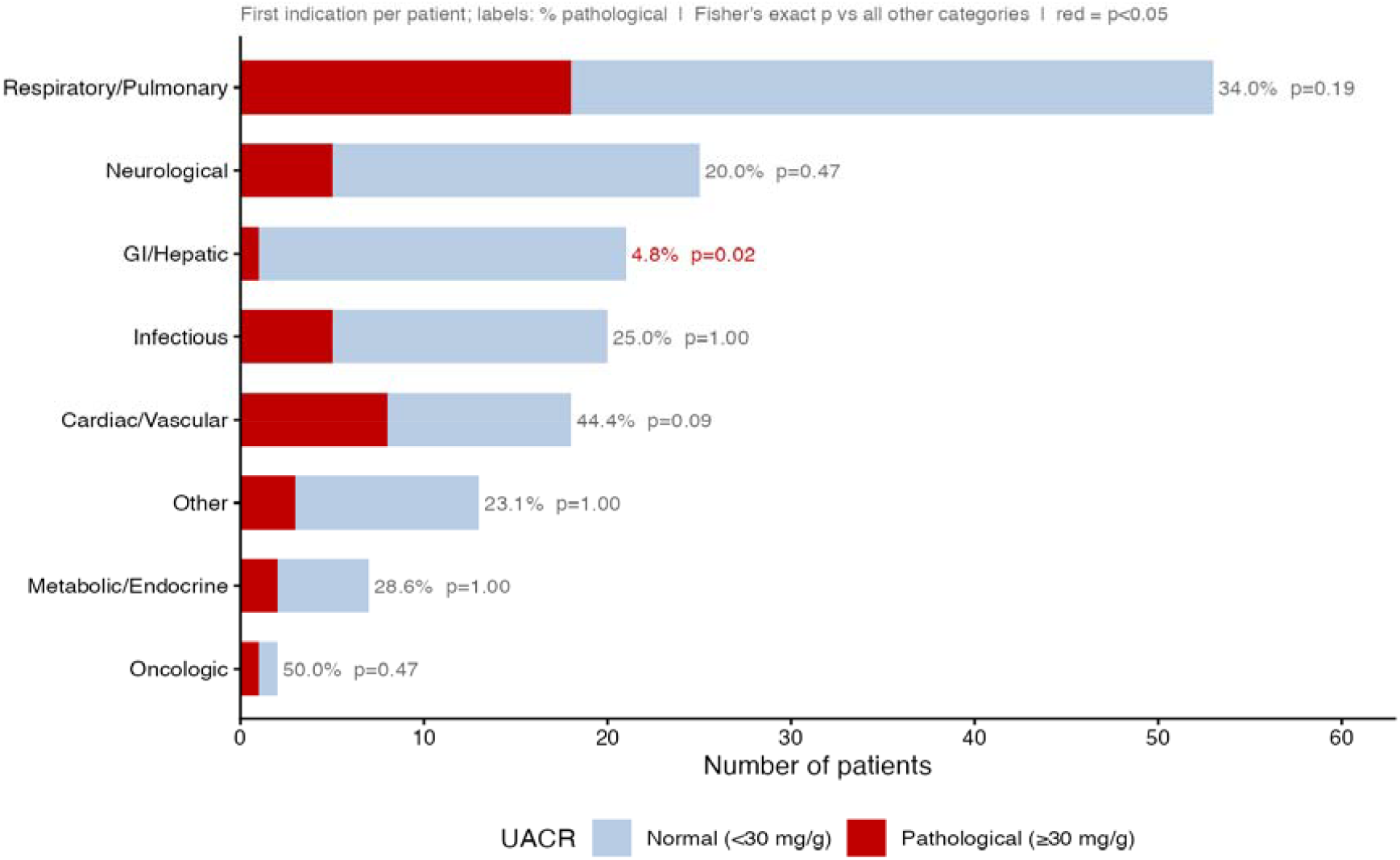
Hospitalization indication categories by UACR status (pooled, n□ = □158). Stacked bars show number of patients with normal (blue) and pathological (red) first UACR. Labels indicate % pathological and Fisher’s exact p-value vs all other categories; red labels = p□<□0.05.

### Limitations

This is a preliminary report from a prospective cohort; the SZMC arm has 4 patients with pending second UACR, and confirmed CKD rates should be regarded as provisional. The ≥90-day KDIGO^[2]^interval criterion has been temporarily relaxed for analysis pending verification of second-UACR dates in the EK cleaned file (two patients with implausible inter-test intervals). Comorbidity data rely on eCRF self-report at enrolment and may underestimate prevalence. The study was not powered to detect differences across indication categories; the indication analysis is exploratory. No adjustment for multiple comparisons has been applied to the indication-level Fisher tests.

## Conclusions

Opportunistic UACR screening during hospitalisation identifies clinically significant and previously unrecognised CKD in approximately 9% of at-risk admitted patients. The screening strategy is feasible, shows consistent results across independent campuses, and identifies a target population for nephrology follow-up. BUN elevation and heart failure are the strongest independent predictors of a positive screen in this cohort.

## Data Availability

All data produced in the present study are available upon reasonable request to the authors

## Notes

### Competing Interest Statement

The authors have declared no competing interest.

### Author Declarations

The Helsinki (ethical) Committee of The Hadassah Medical Organization gave ethical approval for this work.

### Funding

- AstraZenaca (Israel)

